# Correcting for effect modification in the doubly-ranked non-linear Mendelian randomization method

**DOI:** 10.64898/2026.01.22.26344640

**Authors:** Ang Zhou, Haodong Tian, Ashish Patel, Amy Mason, Guoyi Yang, Elina Hyppönen, Stephen Burgess

## Abstract

The doubly-ranked non-linear Mendelian randomization method can yield biased estimates when instrument strength varies across individuals due to gene-environment (GxE) interactions. We propose a simple strategy to mitigate this bias by modelling GxE interactions and removing the fitted GxE component from the exposure before stratification by the doubly-ranked method. In simulations, the proposed GxE correction strategy eliminated GxE-induced bias with null, linear and non-linear exposure-outcome relationships, and it did not introduce bias even when the effect modifier of the IV-exposure association was a confounder or was correlated with a mediator or collider of the exposure-outcome association. In empirical analyses of serum 25(OH)D, BMI, and LDL-C, falsification tests showed bias in the uncorrected doubly-ranked method. Under the selected panel of effect modifiers, the extent of bias attenuation achieved by GxE correction varied by exposures. GxE correction was most effective for LDL-C, with further support from analyses using negative controls (age at recruitment and sex) and coronary artery disease as a positive control. These findings provide proof of principle evidence that our proposed GxE correction strategy can mitigate GxE-induced bias in practice. Where applicable, we recommend implementing this GxE correction strategy as a sensitivity analysis to assess the robustness of findings from the doubly-ranked method.

## Introduction

Mendelian randomization (MR) is an epidemiological approach that uses genetic variants as instruments to estimate the causal effect of an exposure on an outcome [1]. Standard MR assumes a linear exposure-outcome relationship, which may not hold in many biomedical settings where homeostatic mechanisms operate, and adverse effects of an exposure often only emerge below or above certain thresholds. Non-linear MR (NLMR) extends the standard MR framework by allowing the exposure effect to vary across exposure levels, enabling more accurate characterization of the shape of the exposure-outcome relationship. In the stratification-based NLMR, the cohort is first stratified into subgroups with different exposure levels. Localized average causal effect (LACE) estimates are then computed within each subgroup and subsequently modelled [2] to reconstruct the overall exposure-outcome curve.

The stratification step in NLMR is non-trivial. Naïve stratification by exposure level can introduce collider bias, as in an MR setting, the exposure is a common effect of genetic instruments and confounders. An ideal stratifier should capture variation in the exposure while being independent of genetic instruments, thereby preventing collider bias and enabling unbiased LACE estimates [3] (Figure 1A). The doubly-ranked stratification method has recently been proposed to construct such a stratifier [4]. In this approach, participants are first ranked into pre-strata according to their genetic instrument levels, and then ranked within each pre-stratum by their exposure levels. This exposure ranking is then used to stratify the population [4]. If the genetic instrument takes exactly the same value within each pre-stratum, then the exposure ranking reflects variation in exposure levels while remaining independent of the genetic instruments. In practice, values of the genetic instrument will vary within the pre-strata, but this variation is typically small and hence any association between the instrument and the exposure ranking should be minimal.

**Figure 1.**
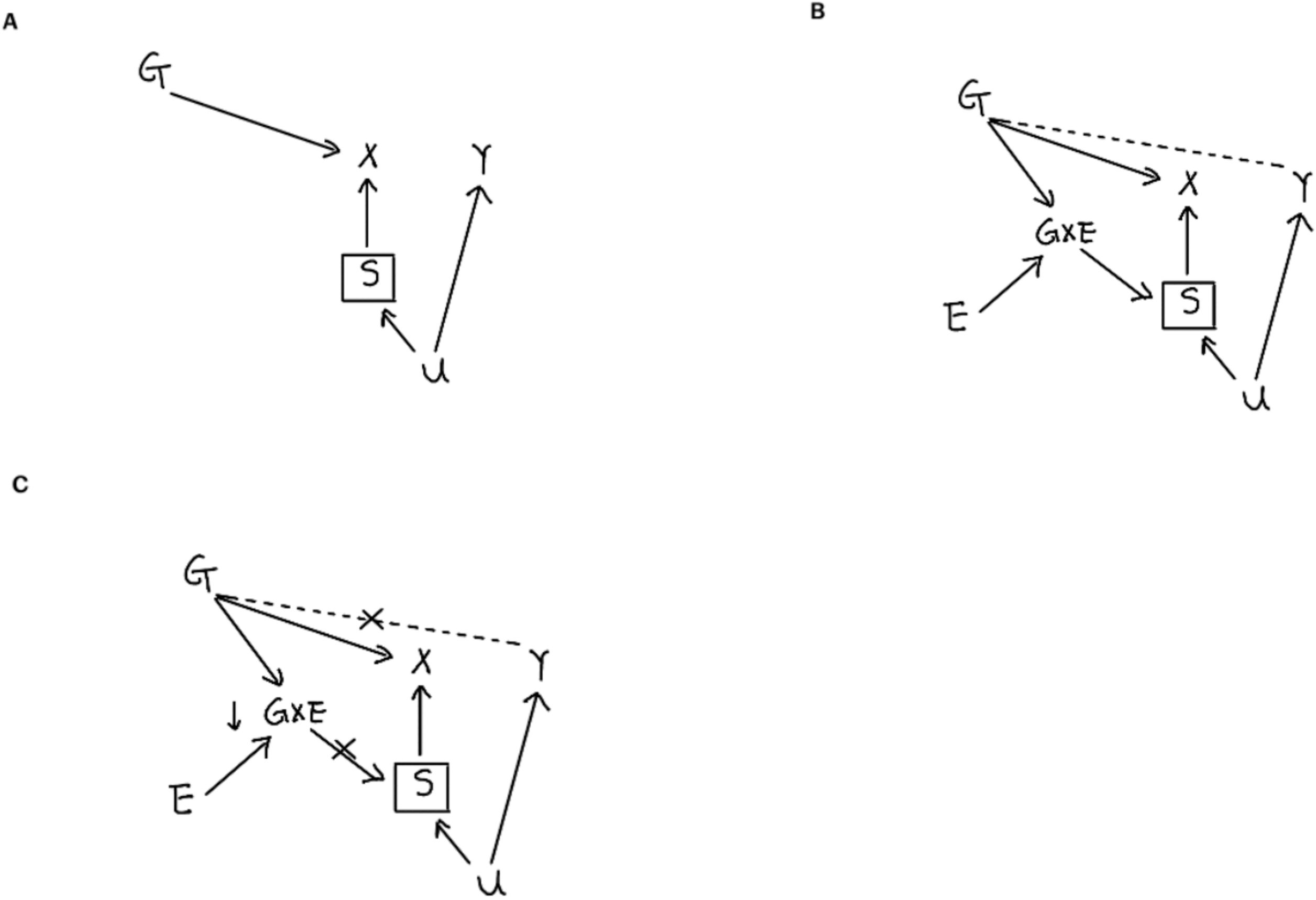
Directed acyclic graphs illustrating how GxE interaction can bias the doubly-ranked method, and how the proposed GxE-correction mitigates this bias. G: genetic instrument; X: exposure; Y: outcome; E: effect modifier of G-X association; U: confounder; GxE: effect modification of the G-X association; S: stratifier. (A) Under the rank-preserving assumption, S is not a function of G; Conditioning on S therefore does not induce bias. (B) If S is akected by a gene-environment interaction (GxE), then it is a common ekect (collider) of G and U. Conditioning on S induces an association between G and U, and hence an association between G and the outcome Y. (C) By removing the dependence of stratification on GxE, S is no longer a collider of G and U. Crossed lines indicate that the GxE-induced pathway is effectively removed, thereby removing this source of bias.

As with all statistical methods, the validity of the doubly-ranked method relies on an underlying assumption: the rank-preserving assumption, which states that the ranking of participants by their exposure values would be the same at all values of the genetic instrument [4]. When this assumption is violated, the stratifier generated by the doubly-ranked method may no longer retain its desirable properties, and stratification may introduce collider bias, leading to biased LACE estimates.

Recent simulation analyses have shown that effect modification of the IV-exposure association, at magnitudes comparable to those observed in practice, can violate the rank-preserving assumption and bias LACE estimates across strata produced by the doubly-ranked method [5] (Figure 1B). This highlights a potential source of bias and limits the applicability of the method, given that gene-environment (GxE) interaction is commonly observed in practice. To address this, in the current study we explored whether residualizing out GxE interactions from the exposure before applying the doubly-ranked method can mitigate such violation and improve its reliability. We first assessed this strategy in simulations where true underlying effect modifiers are known and the GxE interaction can be fully accounted for, and then extended the evaluation to empirical settings where true effect modifiers are unknown and complete GxE correction is unlikely to be achievable. In the latter, we first reproduced previously reported biases in falsification tests for serum 25(OH)D, body mass index (BMI), and low-density lipoprotein cholesterol (LDL-C) [5], and then assessed the extent to which correcting for selected effect modifiers could mitigate these biases.

## Materials and Methods

### Proposed strategy

The doubly-ranked method uses successive rankings: first, individuals are stratified into pre-strata by instrument level; second, within each pre-stratum, they are ranked by exposure to form the final strata [4]. Our proposed strategy modifies the second step by ranking individuals within each pre-stratum according to a GxE-corrected exposure (i.e. the exposure residualized on GxE interactions) rather than the raw exposure. Assuming the effect heterogeneity model is correctly specified, the GxE correction removes exposure variation attributable to GxE interactions, such that the instrument strength does not vary across effect modifier levels. This satisfies the rank-preserving assumption in the presence of known effect modification and, in turn, prevents collider bias (Figure 1C).

It is important to note that the GxE-corrected exposure is used only for the stratification step because the GxE-induced bias that we aim to address arises specifically during stratification (Figure 1B). Once strata are formed, LACE estimates within each stratum are computed using the original exposure, so that the estimand remains the causal effect of the original exposure on the outcome and is consistent with the standard linear MR estimation. There are many ways to model GxE interactions. In this study, we used a standard regression-based interaction model with explicit GxE product terms [6], which is widely used in the applied GxE analyses. Specifically, we regressed the exposure on its corresponding genetic score, potential effect modifiers, and their GxE interaction terms. The GxE-corrected exposure was computed by subtracting the fitted GxE component from the exposure.

### Simulation study

Our simulation study includes three scenarios, each examined under three conditions. The corresponding data generating models (DGM) for all three scenarios and conditions are detailed in Table 1.

- In scenario 1, we assessed the influence of the ‘net’ GxE interaction on the performance of the doubly-ranked method. Here, we define the ‘net’ GxE interaction as the overall interaction between the genetic score and a set of ekect modifiers, obtained by combining the individual GxE interaction terms into a single composite interaction term. A gradient of the ‘net’ GxE interaction was created by varying the correlation between two ekect modifiers, ranging from no ‘net’ GxE interaction when the modifiers were perfectly correlated to the strongest net interaction when they were uncorrelated.
- In scenario 2, we evaluated the performance of our proposed strategy with null, linear, and non-linear exposure-outcome relationships.
- In scenario 3, we examined whether the GxE correction could potentially introduce bias when the ekect modifier of the IV-exposure association also acts as a confounder or is highly correlated with a mediator or collider of the exposure-outcome association.

**Table 1.** Data generating mechanisms for simulation scenarios.

| DGM | DAG |
| --- | --- |
| **Scenario 1A (two uncorrelated effect modifiers, null effect)**<br>$X = 0.3G + 0.3E_1 + 0.2E_2 + 0.1(GxE_1) - 0.1(GxE_2) + 0.3U + e_x$<br>$Y = 0.3U + e_y$<br>$G, U, E_1, E_2, e_x \text{ and } e_y \sim N(2, 1)$<br>$\text{corr}(E_1, E_2) = 0$ | |
| **Scenario 1B (two partially correlated effect modifiers, null effect)**<br>$X = 0.3G + 0.3E_1 + 0.2E_2 + 0.1(GxE_1) - 0.1(GxE_2) + 0.3U + e_x$<br>$Y = 0.3U + e_y$<br>$G, U, E_1, E_2, e_x \text{ and } e_y \sim N(2, 1)$<br>$\text{corr}(E_1, E_2) = 0.5$ | |
| **Scenario 1C (two fully correlated effect modifiers, null effect)**<br>$X = 0.3G + 0.3E_1 + 0.2E_2 + 0.1(GxE_1) - 0.1(GxE_2) + 0.3U + e_x$<br>$Y = 0.3U + e_y$<br>$G, U, E_1, E_2, e_x \text{ and } e_y \sim N(2, 1)$<br>$\text{corr}(E_1, E_2) = 1$ | |
| **Scenario 2A (one effect modifier, null effect)**<br>$X = 0.3G + 0.3E + 0.1(GxE) + U + e_x$<br>$Y = U + e_y$<br>$G, U, E, e_x \text{ and } e_y \sim N(2, 1)$ | |
| **Scenario 2B (one effect modifier, linear effect)**<br>$X = 0.3G + 0.3E + 0.1(GxE) + U + e_x$<br>$Y = 0.5X + U + e_y$<br>$G, U, E, e_x \text{ and } e_y \sim N(2, 1)$ | |
| **Scenario 2C (one effect modifier, quadratic effect)**<br>$X = 0.3G + 0.3E + 0.1(GxE) + U + e_x$<br>$Y = 0.1X^2 + U + e_y$<br>$G, U, E, e_x \text{ and } e_y \sim N(2, 1)$ | |
| **Scenario 3A (one effect modifier that is a confounder, null effect)**<br>$X = 0.3G + 0.3E + 0.1(GxE) + e_x$<br>$Y = E + e_y$<br>$G, U, E, e_x \text{ and } e_y \sim N(2, 1)$ | |
| **Scenario 3B (one effect modifier that affects a mediator, linear effect)**<br>$X = 0.3G + 0.3E + 0.1(GxE) + U + e_x$<br>$M = 0.5X + E + e_m$<br>$Y = M + U + e_y$<br>$G, U, E, M, e_x \text{ and } e_y \sim N(2, 1)$ | |
| **Scenario 3C (one effect modifier that affects a collider, null effect)**<br>$X = 0.3G + 0.3E + 0.1(GxE) + U + e_x$<br>$C = 0.5X + E + 0.5Y + e_c$<br>$Y = U + e_y$<br>$G, U, E, C, e_x \text{ and } e_y \sim N(2, 1)$ | |

Each simulation scenario was repeated 100 times, with a sample size of 100,000 per replicate. DGM: data generating mechanism; DAG: directed acyclic graph

In all scenarios, we performed linear and non-linear MR analyses. Non-linear MR analyses were conducted using the doubly-ranked method. In scenarios 2 and 3, we compared the performance of the doubly-ranked method without and with GxE correction. When applying GxE correction, the GxE-corrected exposure was computed by first fitting a linear model regressing the exposure on the IV, the effect modifier, and their GxE interaction term. The fitted GxE component was then subtracted from the exposure to obtain the GxE-corrected exposure. Each simulation scenario was repeated 100 times, with a sample size of 100,000 per replicate.

### Empirical study

We evaluated the performance of our proposed strategy in the empirical setting using a previously proposed falsification test [5], which involves simulating outcome data and linking it to the real exposure data through a simulated confounder (Figure S1). Since there is no true causal effect of the exposure on the simulated outcome, the falsification test is essentially a negative control analysis. Any deviation of LACE estimates from the null in the falsification test when applying the doubly-ranked method would indicate bias and a potential violation of the rank-preserving assumption [5] for the given IV-exposure association. In our empirical analysis, we considered three exposures: serum 25(OH)D, BMI and LDL-C, for which the corresponding IV-exposure associations in UK Biobank, have been reported to yield biased LACE estimates in the falsification test [5,7].

- **Cohort:** All empirical data used in the falsification test were obtained from the UK Biobank, a prospective cohort of over 500,000 participants aged 37-73 years recruited from 22 assessment centres across the UK between 2006 and 2009 [8]. Analytical sample sizes varied across analyses for serum 25(OH)D (n=294,626) [9], LDL-C (n=286,423) [10] and BMI (n=301,354) [11] because data were drawn from dikerent projects, each with slightly dikerent inclusion criteria and availability of data on exposures, outcomes, and covariates.
- **Exposure measurements:** Serum 25(OH)D concentration (nmol/L) was measured using the LIAISON XL 25(OH)D assay (DiaSorin, Stillwater, USA). Plasma LDL-C (mmol/L) was measured using the Beckman Coulter AU5800 (Beckman Coulter Ltd, UK) by enzymatic protective selection analysis. BMI (kg/m²) was defined as weight (measured to the nearest 0.1 kg) divided by the square of height (measured in whole centimetres using a Seca 202 device).
- **Genetic instruments**: Weighted genetic scores comprising independent, genome-wide significant variants were constructed to instrument each of the three exposures. For 25(OH)D, we included two sets of genetic instruments: the replicated score, consisting of 35 variants with replication evidence in an independent cohort [9], and the focused score consisting of 21 variants restricted to gene regions involved in vitamin D metabolism [12]. We standardized both scores to make results comparable across instrument sets. The genetic score for LDL-C was based on 313 variants [10], and that for BMI on 77 variants [11]. Details of the genetic score construction, including variant selection, weighting, and calculation, are provided in the original publications. As noted above, the associations between the four genetic instruments and their corresponding exposures in UK Biobank have previously been reported to yield biased LACE estimates in the falsification test [5,7].
- **Simulated negative control outcome:** In the falsification test, negative control outcome data were simulated for each exposure according to the data-generating mechanism described previously [5] and depicted in Figure S1. Let X_real_ denote the observed exposure, X_sim_ the simulated exposure, and U_sim_ a simulated confounder. The negative control outcome Y_sim_ was generated as:

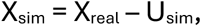

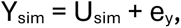

where X_real_ has been scaled to have a mean of 0 and standard deviation of 1, U_sim_ ∼ N(2, 1) and e_y_ ∼ N(2, 1). The simulation was repeated 100 times for each exposure.
- **EEect modifiers and GxE-corrected exposures**: To construct GxE-corrected exposures, we included four core potential ekect modifiers spanning dikerent domains: age at recruitment and sex, representing basic demographics; Townsend deprivation index (TDI), reflecting area-level deprivation and socio-demographics; and frailty index (FI) [13], capturing health status among aging individuals. We additionally included month of blood collection for 25(OH)D and physical activity for BMI, given the known influence of season on 25(OH)D levels [14] and of physical activity on BMI. A higher TDI means a greater degree of deprivation, and a higher FI score indicates greater frailty. Physical activity was measured as the metabolic equivalent task (MET)-minutes/week [15], with higher MET values reflecting higher levels of physical activity. As described earlier, GxE-corrected exposure was computed by first fitting a linear model regressing the exposure on the corresponding genetic score, the selected ekect modifier(s), and their GxE interaction term(s). The fitted GxE component was then subtracted from the exposure to obtain the GxE-corrected exposure. For LDL-C, we considered six GxE corrections: (1) GxAge; (2) GxSex; (3) GxTDI; (4) GxFI; (5) GxE-FO, all first-order GxE interaction terms combined; and (6) GxE-HO, all first-order GxE interaction terms combined plus higher-order GxE interaction terms. For 25(OH)D, we additionally included a GxMonth correction, which was also incorporated into the GxE-FO and GxE-HO specifications for 25(OH)D. For BMI, the additional GxE correction was GxMET (physical activity), which was likewise incorporated into the GxE-FO and GxE-HO specifications for BMI. In total, we evaluated six GxE corrections for LDL-C and seven for each of 25(OH)D and BMI. Regression models corresponding to each GxE correction are presented in Table S1.
- **Replication using empirical negative/positive controls:** For IV-exposure associations where GxE correction showed clear improvements, we carried forward the two best-performing GxE correction specifications from the falsification test and evaluated them further using empirical negative control outcomes, age at recruitment and sex. For the LDL-C genetic score, we additionally evaluated the top-performing GxE correction using a positive control outcome, coronary artery disease (CAD, N_case_ = 19,009) [16]. All these analyses were adjusted for age at recruitment (except when age was the outcome), sex (except when sex was the outcome), assessment centres, and top 40 genetic principal components to account for population stratification.
- **Statistical analyses:** Non-linear MR analyses were performed using the doubly-ranked method, applied to both the original and GxE-corrected exposures. Where appropriate, linear MR analyses were also conducted. The non-linear and linear MR analyses were carried out using the SUMnlmr R package [17] and MendelianRandomization R package (version 0.10.0) [18], respectively. The doubly-ranked method that accommodates GxE-corrected exposures has been implemented within the create_nlmr_summary function in the SUMnlmr package.

## Results

### ‘Net’ GxE interaction matters

Figure 2 shows the result from simulation scenario 1, which assessed the influence of the ‘net’ GxE interaction effect on the performance of the doubly-ranked method. A gradient of the ‘net’ GxE interaction effect was created by varying the correlation between two effect modifiers. Deviation of the median LACE estimates from the null indicates bias. The largest deviation was observed when the modifiers were uncorrelated (representing the highest ‘net’ interaction), moderate deviation when they were partially correlated (intermediate ‘net’ interaction), and no deviation when they were fully correlated (no ‘net’ interaction, Figure 2).

**Figure 2.**
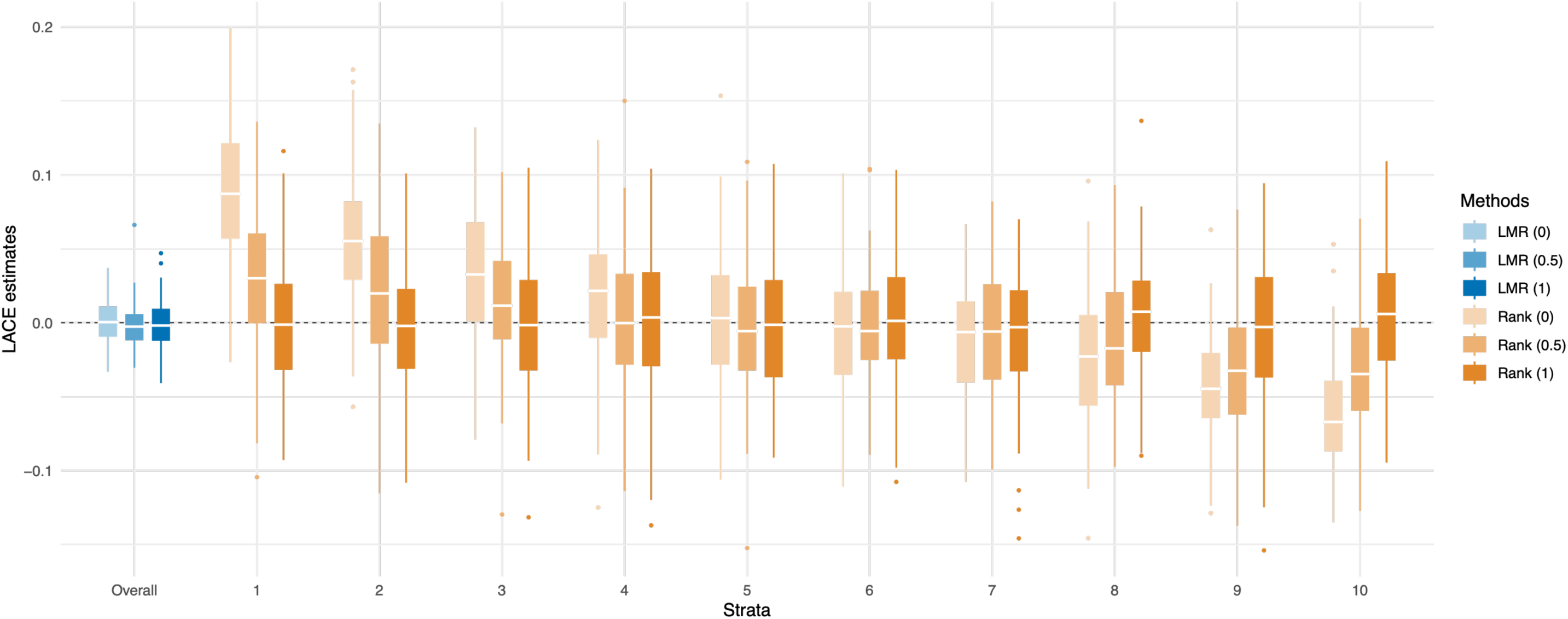
Results for simulation scenario 1 with the net GxE interaction gradient induced by varying the correlation between two effect modifiers at 0 (A), 0.5 (B) and 1.0 (C). Simulation was repeated 100 times. Boxplots represent the distribution of estimates across 100 replicates. The box displays the lower quartile, median, and upper quartile; whiskers extend to the minimum and maximum values within 1.5 x interquartile range from the lower and upper quartiles. Estimates outside this range are shown as individual points. The dotted line indicates the null effect. LACE estimate: localized average causal effect estimate; LMR: linear MR; Rank: doubly-ranked method.

### GxE correction can mitigate bias

Figure 3 presents the results from simulation scenario 2, which evaluated the performance of our proposed strategy with null, linear, and non-linear exposure-outcome relationships. Results for the null exposure-outcome relationship are shown in Figure 3A. Deviation of the median LACE estimates from the null indicates bias. When the original exposure was used with no GxE correction, deviations of the median LACE estimates from the null were observed across strata (Figure 3A). In contrast, after applying our proposed GxE-correction strategy, the median LACE estimates closely aligned with the null across strata (Figure 3A). Similar bias neutralizing effects of our proposed GxE-correction strategy were also observed under both linear (Figure 3B) and non-linear (Figure 3C) exposure-outcome relationships.

**Figure 3.**
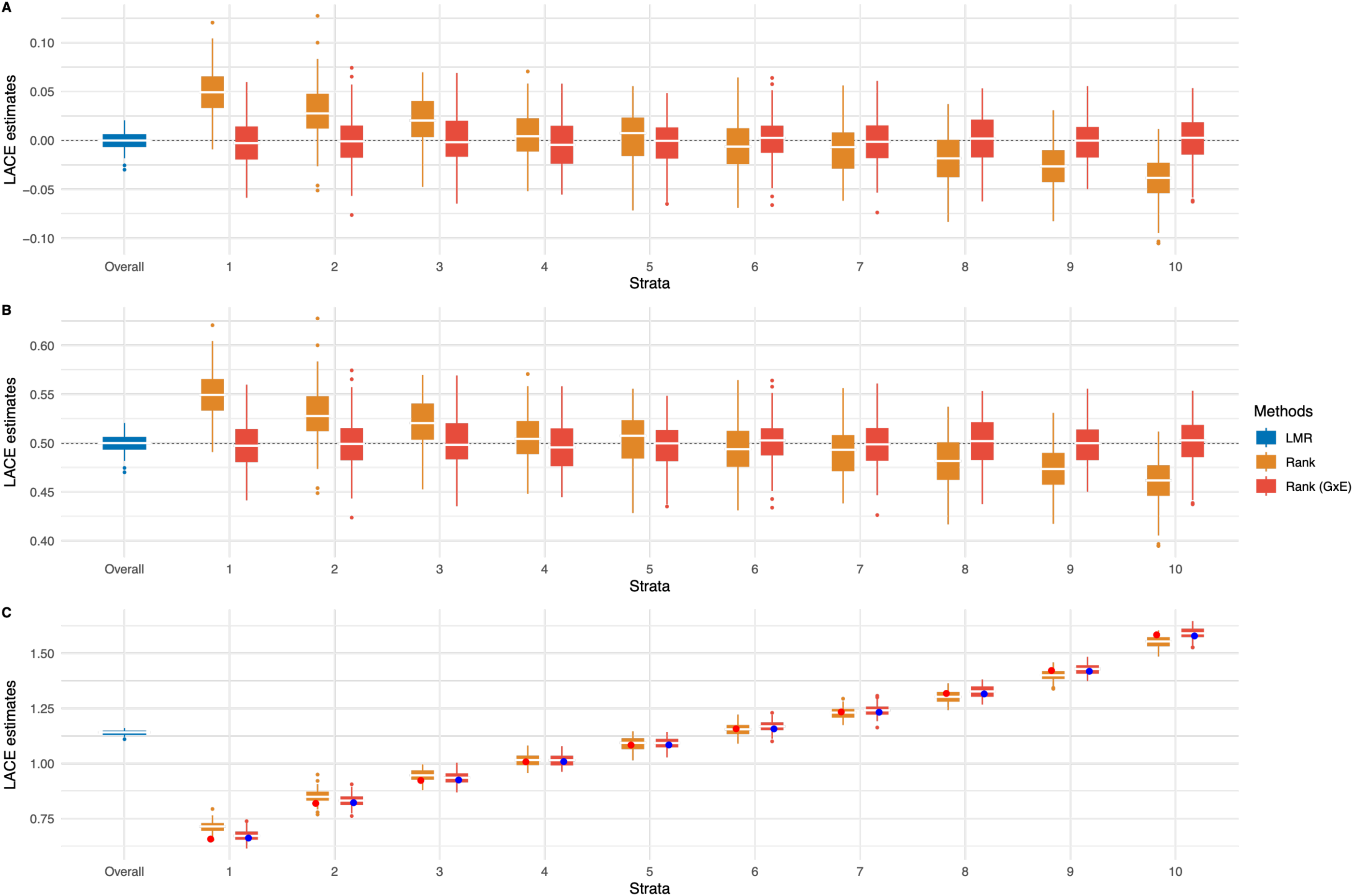
Results for simulation scenario 2 comparing the doubly-ranked method with and without GxE correction under null (A), linear (B), and non-linear (C) exposure-outcome relationships. Simulation was repeated 100 times. Boxplots represent the distribution of estimates across 100 replicates. The box displays the lower quartile, median, and upper quartile; whiskers extend to the minimum and maximum values within 1.5 x interquartile range from the lower and upper quartiles. Estimates outside this range are shown as individual points. In panel C, red and blue points represent the target causal effect in each stratum, calculated as the median across 100 replications of the slope of the true exposure-outcome curve evaluated at that stratum’s mean exposure level. The dotted line indicates the target effect. LACE estimate: localized average causal effect estimate. LMR: linear MR; Rank: doubly-ranked method without GxE correction; Rank (GxE): doubly-ranked method with GxE correction.

### GxE correction itself does not introduce bias

Figure 4 illustrates the results from simulation scenario 3, where we examined whether the GxE correction could introduce bias when the effect modifier also acts as a confounder or is highly correlated with a mediator or collider of the exposure-outcome association. Across all three conditions, there was no evidence that the GxE correction introduced bias into the LACE estimates across strata (Figure 4).

**Figure 4.**
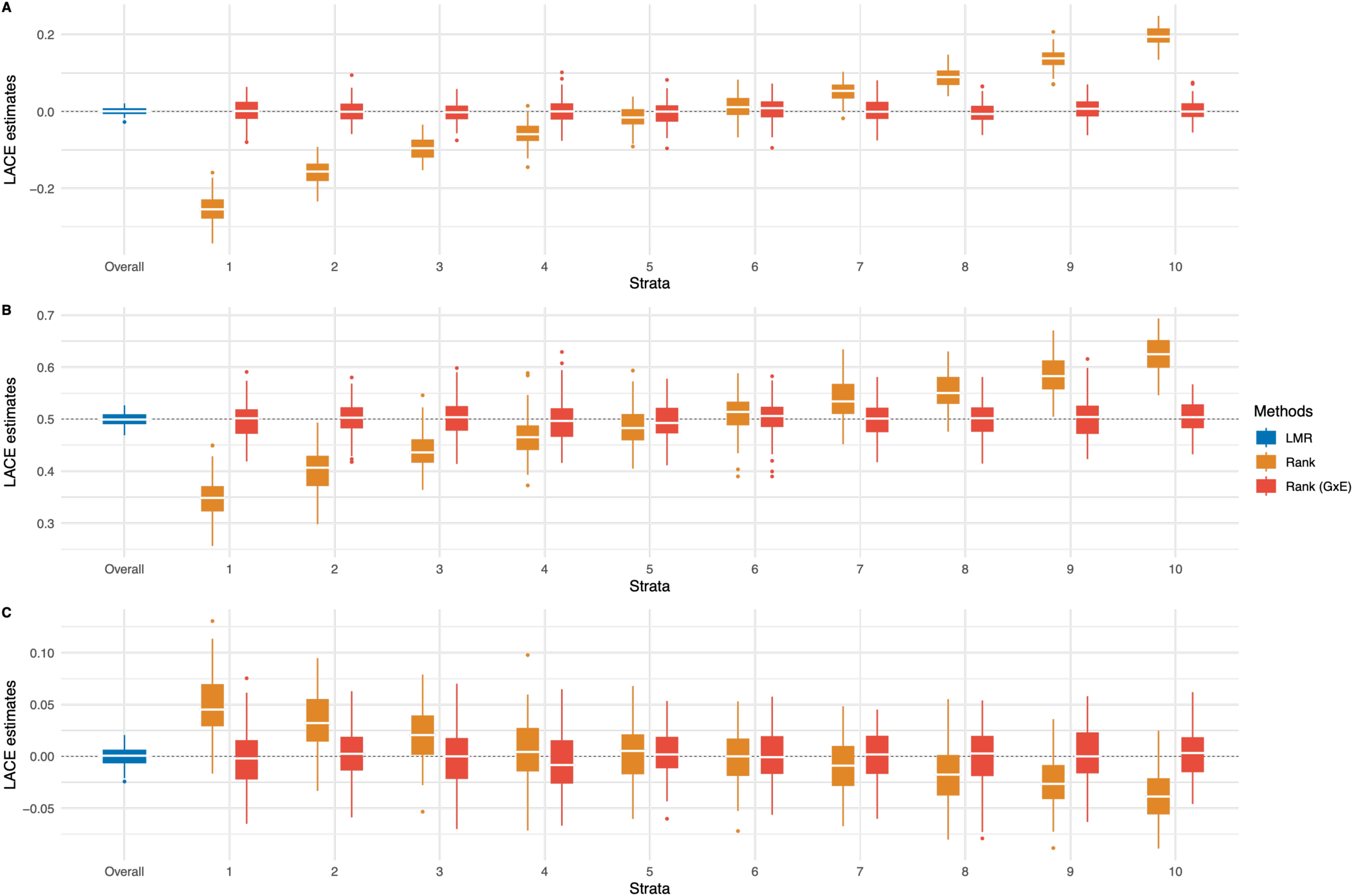
Results for simulation scenario 3 comparing the doubly-ranked method without and GxE correction when the effect modifier is a confounder (A), or is highly correlated with a mediator (B) or collider (C) of the exposure-outcome relationship. Simulation was repeated 100 times. Boxplots represent the distribution of estimates across 100 replicates. The box displays the lower quartile, median, and upper quartile; whiskers extend to the minimum and maximum values within 1.5 x interquartile range from the lower and upper quartiles. Estimates outside this range are shown as individual points. The dotted line indicates the target effect. LACE estimate: Localized average causal effect estimate. LMR: linear MR; Rank: doubly-ranked method without GxE correction; Rank (GxE): doubly-ranked method with GxE correction.

### GxE correction can mitigate bias in empirical settings when relevant effect modifiers can be identified

In our empirical analyses, we considered three exposures (25(OH)D, BMI, and LDL-C) and used four genetic instruments: two instruments for 25(OH)D (the replicated and focused scores) and one instrument each for BMI and LDL-C. We first examined effect modification of the IV-exposure association by six effect modifiers: four core modifiers (age at recruitment, sex, TDI, and FI) across all IV-exposure associations, with month of blood collection additionally considered for the IV-25(OH)D association and physical activity additionally considered for the IV-BMI association. Evidence of effect modification was observed for all IV-exposure associations across all effect modifiers, except for the IV-LDL-C association with TDI (Figure 5). For 25(OH)D, effect modification patterns were similar for the replicated and focused scores, although they appeared stronger for the focused score (Figure 5A and 5B). Month of blood collection was the strongest effect modifiers for the IV-25(OH)D association (P_interaction_ = 2 ×10^-68^ for the replicated score and 9 ×10^-183^ for the focused score), FI was the strongest effect modifier for the IV-BMI association (P_interaction_ = 4 ×10^-53^), and age was the strongest effect modifier for the IV-LDL-C associations (P_interaction_ = 7 ×10^-134^) (Figure 5).

**Figure 5.**
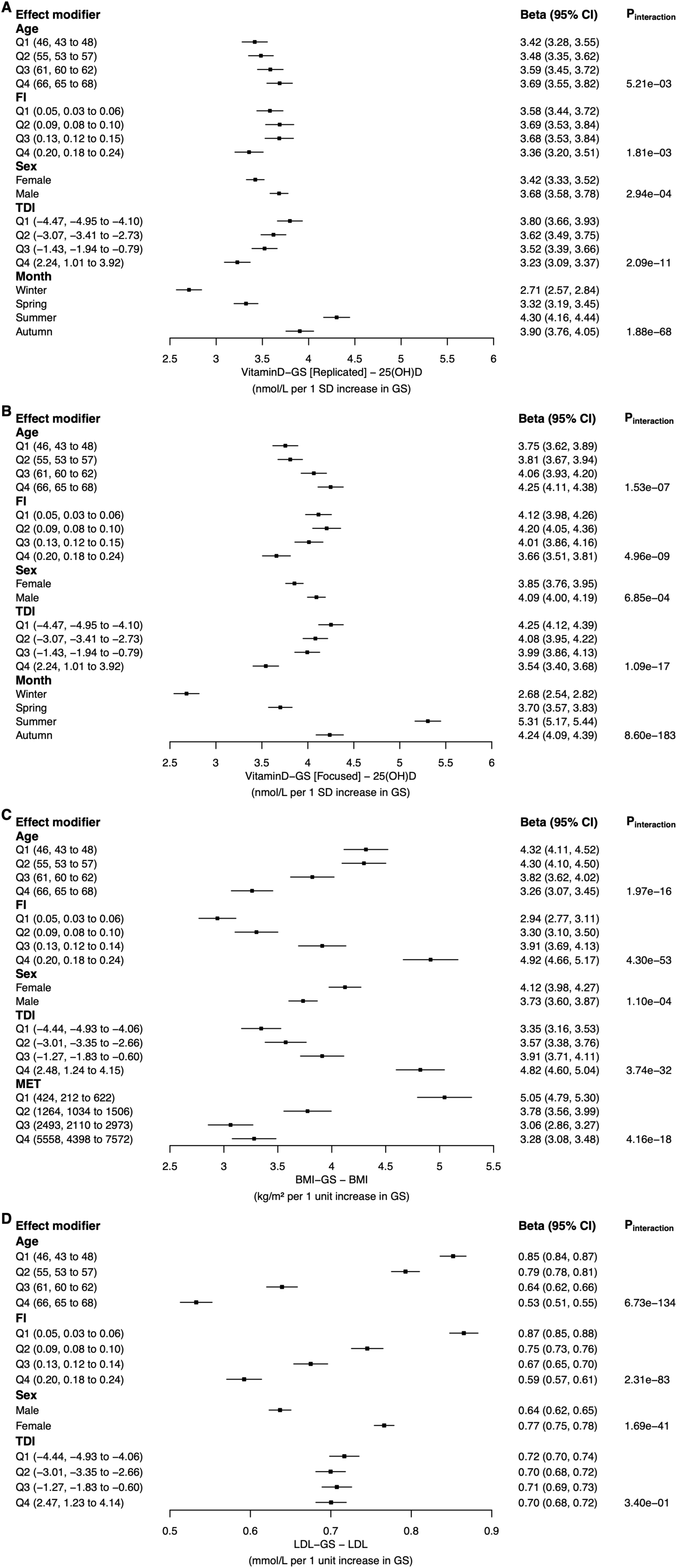
Effect modification of the genetic score-exposure association for 25(OH)D using the replicated score (A), 25(OH)D using the focused score (B), BMI (C), and LDL-C (D), by age at recruitment, frailty index, sex, Townsend deprivation index, month of blood collection and physical activity measured by metabolic equivalent task (MET). For age, Frailty index, Townsend deprivation index and MET score, results are presented by quartiles. Values in brackets for each quartile indicate the median and the 25th and 75th percentiles of the effect modifier, with Q1 being the lowest and Q4 the highest quartile. Month of blood collection was grouped into seasons. TDI: Townsend deprivation index; FI: frailty index; MET: metabolic equivalent task; SD: standard deviation. P_interaction_: p value for the GxE interaction term(s) from covariate-adjusted linear regressions of exposure on G, E, and GxE. Error bars are 95% CIs.

We evaluated our proposed GxE correction strategy in the falsification test, with seven GxE correction specifications for 25(OH)D and BMI, and six for LDL-C. Figure 6 illustrates the performance of the two best-performing GxE specifications (ranked by mean-squared error, Figure 7) for each IV-exposure association, with results for all specifications shown in Figure S2. In this setting, deviations of the median LACE estimates from the null indicate bias. In absence of GxE correction, deviations were observed for all IV-exposure associations (Figure 6), consistent with previous findings [5,7]. When applying GxE correction, bias attenuation was most pronounced for the IV-LDL-C association and was evident across all strata (Figures 6D and 7). More modest attenuation was observed for the IV-25(OH)D association, with bias reduction primarily confined to the lower strata (Figures 6A, 6B and 7), whereas little evidence of attenuation was seen for the IV-BMI association (Figures 6C and 7). For IV-25(OH)D, the focused score appeared to benefit more from GxE correction than the replicated score (Figures 6A, 6B and 7).

**Figure 6.**
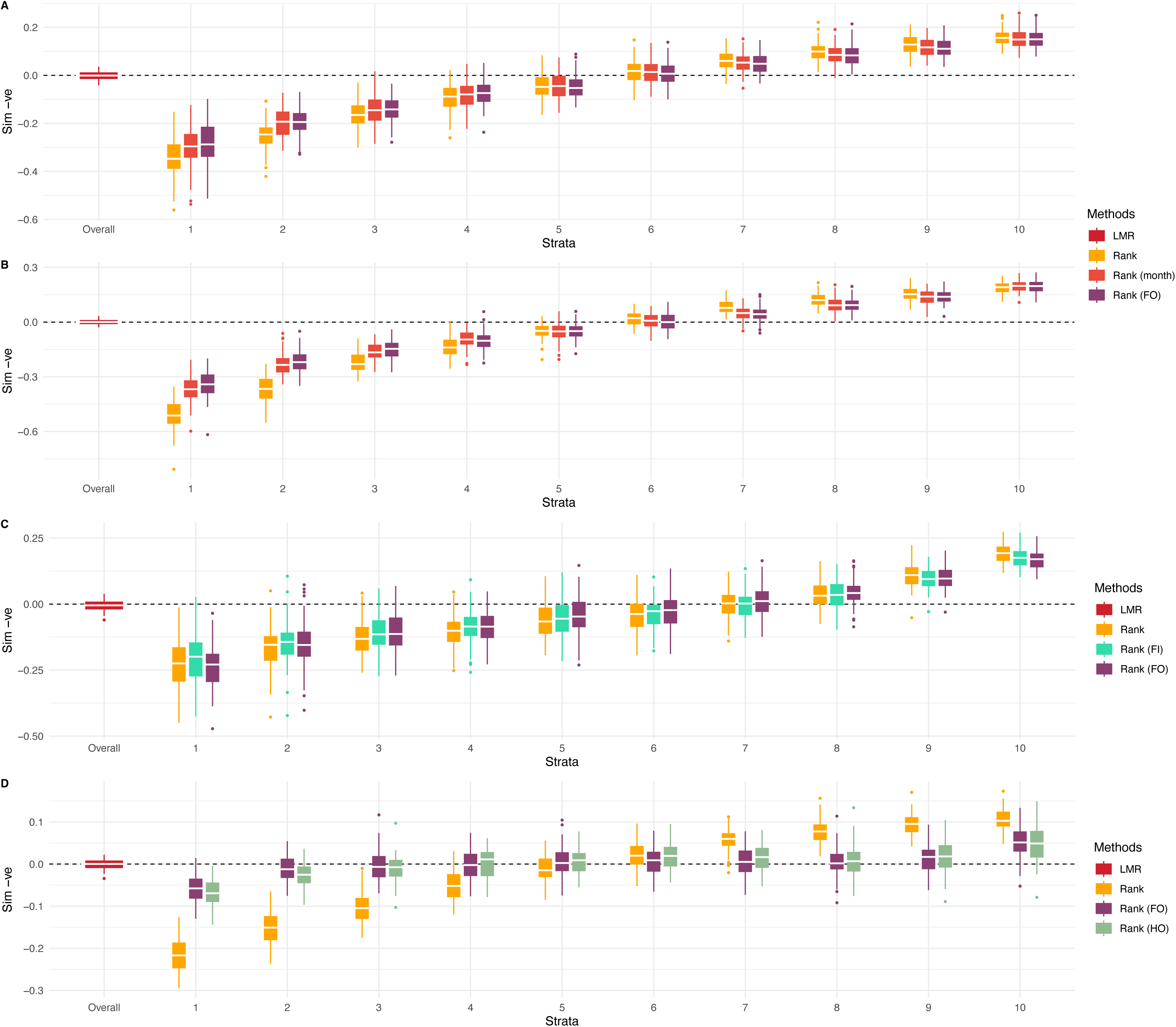
Falsification tests for 25(OH)D using the replicated score (A), 25(OH)D using the focused score (B), BMI (C) and LDL-C (D) comparing the doubly-ranked method without GxE correction with the two best-performing GxE correction specifications. The dotted line indicates the null effect. For each exposure, negative control outcome data were simulated 100 times. Boxplots represent the distribution of estimates across 100 replicates. The box displays the lower quartile, median, and upper quartile; whiskers extend to the minimum and maximum values within 1.5 x interquartile range from the lower and upper quartiles. Estimates outside this range are shown as individual points. LMR: linear MR; Rank: doubly-ranked method applied to the original exposure (no GxE correction); Rank(month) or Rank(FI): doubly-ranked method applied to a GxE-corrected exposure using a single effect modifier, either month of blood collection (month) or frailty index (FI); Rank(FO): doubly-ranked method applied to an exposure corrected using all first-order GxE interaction terms; Rank(HO): doubly-ranked method applied to an exposure corrected using the first-order GxE interaction terms plus higher-order interaction terms; FI: frailty index; Sim -ve: Simulated negative control outcome.

**Figure 7.**
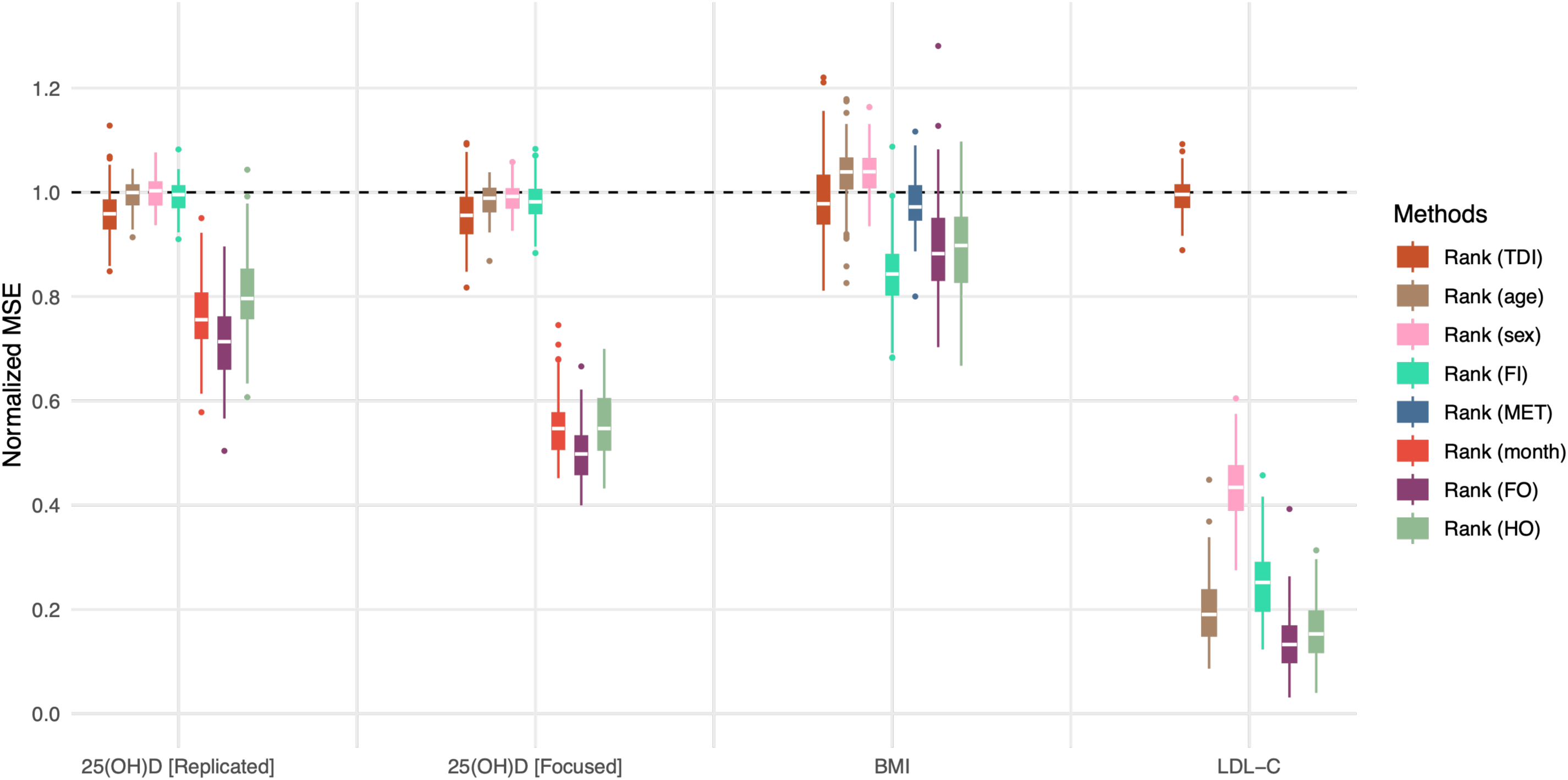
Normalized mean-squared error of LACE estimates from falsification tests, comparing the doubly-ranked method with different GxE correction specifications for 25(OH)D, BMI and LDL-C. For each exposure, negative control outcome data were simulated 100 times. In each replicate, the mean-squared error (MSE) for each GxE correction was calculated as the mean of the squared LACE estimates across the 10 strata and then normalized by dividing by the corresponding MSE from the doubly-ranked method without GxE correction. A normalized MSE of 1 indicates identical MSE to the doubly-ranked method without GxE correction; values < 1 indicate lower MSE (better performance), and values > 1 indicate higher MSE (worse performance). Boxplots show the distribution of normalized MSE values across the 100 replicates. The box displays the lower quartile, median, and upper quartile; whiskers extend to the minimum and maximum values within 1.5 x interquartile range from the lower and upper quartiles. Normalized MSEs outside this range are shown as individual points. 25(OH)D [Replicated]: analyses of 25(OH)D using the replicated score as the genetic instrument; 25(OH)D [Focused]: analyses of 25(OH)D using the focused score as the genetic instrument; Rank: doubly-ranked method applied to the original exposure (no GxE correction); Rank(TDI), Rank(age), Rank(sex), Rank(FI), Rank(month) or Rank(MET): doubly-ranked method applied to a GxE-corrected exposure using a single effect modifier, namely Townsend deprivation index (TDI), age, sex, frailty index (FI), month of blood collection (month) or physical activity measured by metabolic equivalent task (MET), respectively; Rank(FO): doubly-ranked method applied to an exposure corrected using all first-order GxE interaction terms; Rank(HO): doubly-ranked method applied to an exposure corrected using the first-order GxE interaction terms plus higher-order interaction terms; TDI: Townsend deprivation index; FI: frailty index; MSE: mean-squared error; MET: metabolic equivalent task; LACE estimate: localized average causal effect estimate.

For 25(OH)D and LDL-C, where GxE corrections showed clear improvements in the falsification test (Figure 7), we further evaluated the top two performing correction specifications for each exposure using the negative control outcomes of age at recruitment and sex. As expected, the linear MR provided no evidence that genetically predicted 25(OH)D or LDL-C was associated with age or sex (Figure S3, S4 and 10). For 25(OH)D, although bias was evident with the simulated negative control in the falsification test (Figure 6A and 6B), it was less pronounced in the empirical negative control analysis using age and sex. Specifically, using the replicated score, 95% confidence intervals for LACE estimates overlapped the null linear MR estimates across all strata for both age and sex (Figure S3). Using the focused score, results were broadly similar, with only one clear departure of the LACE estimate from the null linear MR estimate, observed in stratum 1 for age at recruitment (Figure S4). After applying the GxE correction, the results remained very similar to those obtained without correction (Figures S3 and S4), possibly reflecting minimal bias in the uncorrected analysis.

For LDL-C, LACE estimates from the uncorrected doubly-ranked method showed clear departures from the null linear MR estimates. These departures were evident across most strata for age, whereas for sex they were largely confined to the higher strata. GxE correction markedly reduced the observed bias (Figure 8A and 8B), particularly for age, with substantially lower mean-squared errors compared to no GxE correction (Figure S5). We additionally evaluated the GxE correction strategy using CAD as a positive control outcome. With the GxE correction applied, LACE estimates across strata became less heterogeneous and more closely aligned with the linear MR estimates (Figure 8C), resulting in much lower mean-squared errors compared to the analysis without GxE correction (Figure S5).

**Figure 8.**
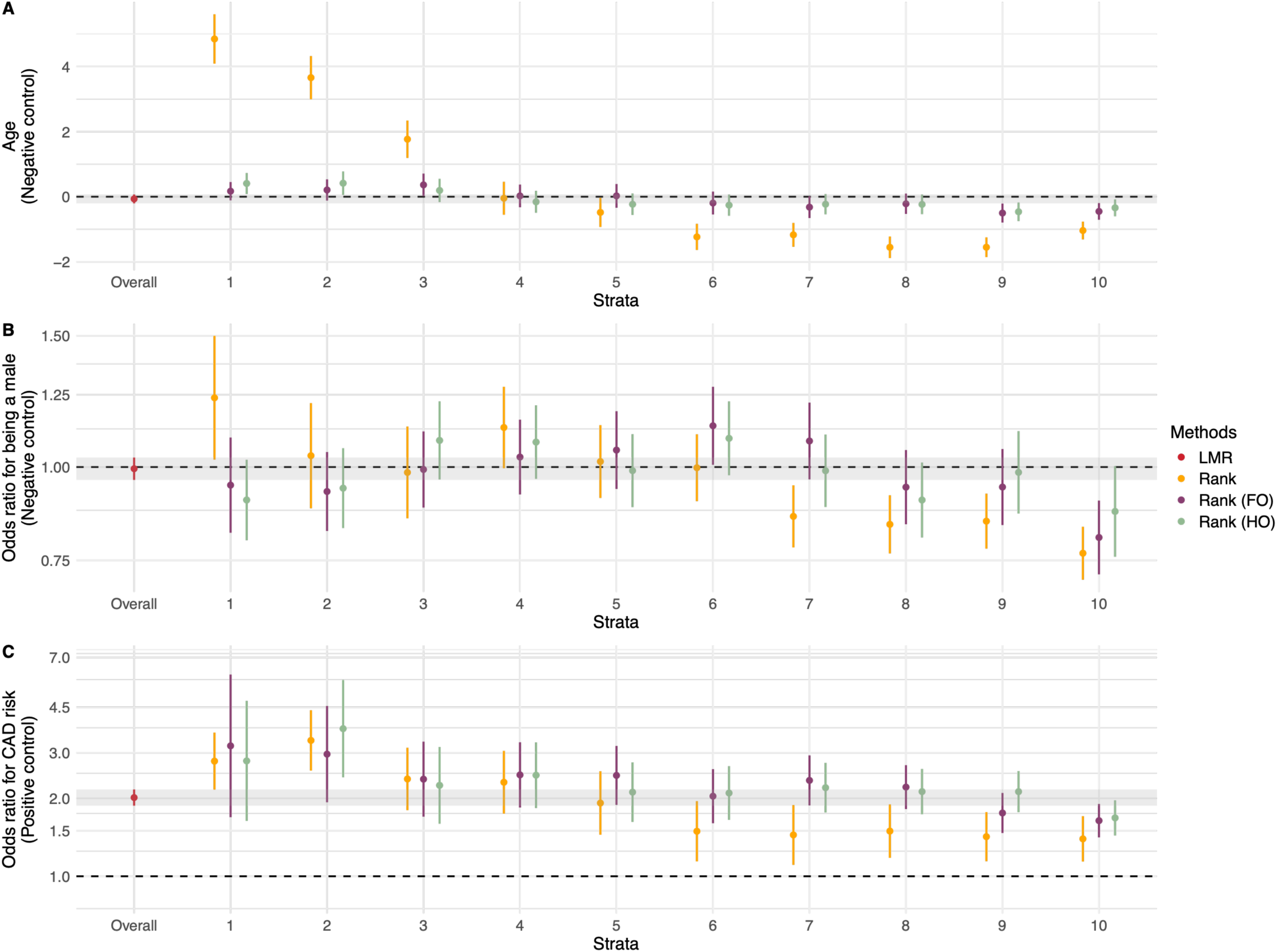
Association of genetically predicted LDL-C with age at recruitment (A), sex (B), and CAD risk (C), comparing the doubly-ranked method without GxE correction with the two best-performing GxE correction specifications. The shaded area shows the 95% CI for the linear MR estimate. The dotted line indicates the null effect. LMR: linear MR; Rank: doubly-ranked method applied to the original LDL-C (no GxE correction); Rank(FO)^1^: doubly-ranked method applied to LDL-C corrected using all first-order GxE interaction terms; Rank(HO)^1^: doubly-ranked method applied to LDL-C corrected using the first-order GxE interaction terms plus higher-order interaction terms; CAD: Coronary artery disease. Error bars are 95% confidence intervals. When computing LACE estimates across strata, all models were adjusted for age (except when age was the outcome), sex (except when sex was the outcome), assessment centres, and top 40 genetic PCs.^1^Given the absence of evidence for GxTDI interaction and the negligible impact of GxTDI correction, TDI was excluded from the GxE-FO and GxE-HO correction models.

## Discussion

Gene-environment (GxE) interactions can violate the rank-preserving assumption and bias estimates from the doubly-ranked method in the presence of confounding [5]. To mitigate this bias, in the current study we propose a simple GxE correction strategy and evaluate its performance using both simulations and empirical data. Simulation analyses illustrate that our proposed strategy performs well in theory when the effect modifiers are known and the GxE interaction can be fully accounted for. Empirical analyses of LDL-C further demonstrate the feasibility of our approach in practice and show that it can also effectively mitigate bias in empirical settings, where complete GxE correction is unlikely to be achievable. Where applicable, we recommend using the proposed GxE correction strategy as a sensitivity analysis to assess the robustness of findings from the doubly-ranked method.

In practice, gene-environment interactions can be complex. Multiple effect modifiers may act simultaneously, each interacting with the genetic predictor of the exposure in different directions and with varying strengths. In our simulation, we saw that the magnitude of bias is determined by the combined (net) interaction rather than the individual interaction effect of each modifier. This suggests that it is the overall ‘net’ interaction landscape that governs the extent of bias in the doubly-ranked method. Therefore, understanding and accounting for the ‘net’ GxE interaction holds the key to mitigating such bias.

Our proposed GxE correction strategy targets the ‘net’ GxE interaction by modelling it and removing it from the exposure. The resulting GxE-corrected exposure is then used in the doubly-ranked method. In our simulation analyses, where the relevant effect modifiers are known and the ‘net’ GxE interaction can be fully accounted for, the proposed GxE correction strategy can completely attenuate the bias due to GxE interaction. In practice, however, uncertainty in the identification and measurement of relevant effect modifiers, as well as in understanding how they interact with the genetic instrument, makes modelling the ‘net’ GxE interaction landscape challenging. As a result, the effectiveness of the GxE correction strategy will depend on how well the underlying ‘net’ GxE interaction landscape can be modelled and accounted for. In our analyses of three exposures, GxE correction was most effective for LDL-C, more modest for 25(OH)D, and showed little impact for BMI. Responsiveness to GxE correction also varied by instrument choice within the same exposure, as illustrated for 25(OH)D, where stronger improvements were seen for the focused score than for the replicated score. Despite heterogeneity in performance across IV-exposure associations, the clear improvement for LDL-C nevertheless demonstrates the feasibility of the GxE correction approach in practice, where complete GxE correction is unlikely to be achievable. Although residual ‘net’ GxE interaction may still exist in the analysis for LDL-C, the expected null results from the empirical negative control outcome analyses with age and sex provide reassurance that its impact may be unlikely to materially affect the findings from the doubly-ranked method.

For LDL-C, we also examined the association between genetically predicted LDL-C and CAD risk as a positive control. Meta-analyses of randomized controlled trials (RCTs) have shown strong, broadly linear effects of LDL-C lowering on CAD risk, with slightly larger effects of more intensive therapy among individuals with higher baseline LDL-C [19–21]. Findings from the doubly-ranked method, both without and with GxE correction, are broadly consistent with RCT findings, showing that higher genetically predicted LDL-C is associated with greater CAD risk. With GxE correction, the stratum-specific causal estimates appear to be more consistent across strata and more closely aligned with the overall linear MR estimate than without GxE correction.

It is important to note that in our empirical analyses, we considered only a limited panel of effect modifiers, most of which (4 of 6) were shared across exposures, and applied a largely ‘one-size-fits-all’ set of GxE correction specifications across different IV-exposure associations. Consequently, the varying degrees of responsiveness to GxE correction across IV-exposure associations do not necessarily imply that the effectiveness of the strategy differs by exposure. Rather, this pattern more likely reflects differences in how well the selected modifiers capture the relevant GxE interaction landscape underlying each IV-exposure association. For exposures such as BMI in our analyses, where little improvement was observed, incorporating additional relevant effect modifiers and adopting more refined GxE modelling in future work could enhance the performance of the GxE correction. Our empirical analyses were intended as a proof of principle demonstration that the GxE correction strategy can mitigate the GxE-induced bias observed in the doubly-ranked method in practice. More refined GxE modelling and hence more nuanced GxE correction tailored to each IV-exposure association is beyond the scope of the current study, but would be an important step in applied analysis.

Given that our GxE correction strategy can mitigate bias in both simulation and empirical settings without introducing additional bias, we recommend using it as a sensitivity analysis to assess the robustness of findings from the doubly-ranked method. Consistent results would strengthen confidence in the findings whereas discrepancy should prompt careful evaluation of their reliability. To implement the GxE correction strategy in the absence of substantive knowledge to guide the modelling, we recommend using the falsification test to inform the modelling of the GxE interaction and selecting the specification that leads to the greatest attenuation of bias in the falsification test. Where possible, the selected GxE correction specification should be further validated using empirical negative and positive control outcomes, as illustrated in our analysis of LDL-C.

Our proposed strategy has some limitations. First, it relies on correct specification of the GxE correction model, which, as noted earlier, is rarely attainable in practice. As a result, the correction may be insufficient. More nuanced modelling of GxE interactions may help address this limitation. For example, in our empirical analyses, we have only considered a limited set of effect modifiers and employed a simple linear framework to model the ‘net’ GxE interaction landscape. Future work could extend our approach by incorporating a systematic screening process for effect modifier selection and exploring more flexible modelling approaches or machine learning techniques to better capture the ‘net’ GxE interaction landscape, thereby minimizing any residual GxE interaction and the resulting bias. Secondly, since the ‘net’ GxE interaction model is selected in a data-driven manner, overfitting is possible. As a result, there is a risk of over-adjustment if an overfitted GxE component removed from exposure inadvertently captures variation related to confounders, mediators, or colliders in the exposure-outcome relationship, thereby introducing bias. Nevertheless, our simulation analyses show that even when an effect modifier is related to a confounder, mediator, or collider in the exposure-outcome relationship, GxE correction does not introduce additional bias. This suggests that the data-driven GxE interaction modelling process is unlikely to pose a major concern. Finally, where possible, the GxE correction strategy should be validated using negative and positive control outcome analyses, which can detect and contextualize biases arising from residual ‘net’ GxE interaction or from over-adjustment.

Finally, we outline considerations for reporting findings from the doubly-ranked method in light of potential GxE-induced bias:

- Falsification tests [5] can be used to probe for potential GxE-induced bias for a given IV-exposure association. Where such bias is suggested, the GxE correction strategy proposed in the current study can be applied as a sensitivity analysis to assess the robustness of findings from the doubly-ranked method. If the GxE correction is not possible or appears insukicient because relevant ekect modifiers are unavailable or largely missing in the dataset, results from the doubly-ranked method should be presented alongside the falsification test and negative control analyses to make uncertainties explicit and to support cautious interpretation. Where possible, inclusion of positive control outcomes can provide additional context, although suitable positive controls may not always be available.
- It is important to note that complete elimination of bias is rarely possible in practice, and the presence of bias does not necessarily imply that bias will materially alter the substantive conclusion. Therefore, although falsification tests and negative control analyses may indicate potential GxE-induced bias, such evidence should not be interpreted as definitive proof that the main conclusions would change. In particular, the magnitude and pattern of bias detected by falsification tests depend on the simulated confounding structure, which may not fully reflect the real-world context under investigation. While negative control outcomes can help contextualize the bias, commonly used controls such as age and sex are typically already adjusted for in MR analyses, and the extent to which such adjustment mitigates any GxE-induced bias remains unclear.
- Additional methodological developments are certainly needed to better address GxE-induced bias and to evaluate the extent to which such bias may materially akect substantive conclusions. Nonetheless, unless there is strong evidence that GxE-induced bias is large enough to change the main interpretation, we believe that reporting findings from the doubly-ranked method remains valuable. These results provide one source of evidence within a broader landscape of epidemiological analyses, enabling triangulation across approaches and ultimately contributing to a more accurate characterization of the shape of the exposure-outcome relationship.

## Conclusion

In summary, we have proposed a simple GxE correction strategy to address bias arising from heterogeneity in the instrument strength due to GxE interactions. Through simulation and empirical analyses, we demonstrate that this strategy can be effective and feasible in practice. Where applicable, we recommend using it as a sensitivity analysis to assess the robustness of findings from the doubly-ranked method.

## Data Availability

Data are available from UK Biobank for researchers who meet the criteria and gain approvals to access the research database from the UK Biobank access management committee at the University of Oxford.

## Author contributions

A.Z. and S.B. conceived of the study. A.Z. analyzed the data with advice from E.H. and S.B.. A.Z. wrote the first draft. All authors interpreted the results, drafted and revised the manuscript critically for important intellectual content, and read and approved the final manuscript.

## Consent to participate

Informed consent was obtained from all participants included in the study.

## Ethics approval

The present study was conducted under UK Biobank application number 20175 and 98032. The UK Biobank study was approved by the National Information Governance Board for Health and Social Care and North West Multicentre Research Ethics Committee (11/NW/ 0382).

## Access to code and data

Data are available from UK Biobank for researchers who meet the criteria and gain approvals to access the research database from the UK Biobank access management committee at the University of Oxford. Analysis code will be available upon publication.

