## Supplementary Materials for "Correcting for effect modification in the doubly-ranked non-linear Mendelian randomization method"

**Table S1 GxE correction specification for each exposure in the falsification test**

| GxE correction | Model specification | GxE-corrected exposure |
|---|---|---|
| GxAge | $Exp = \beta_0 + \beta_1 G + \beta_2 Age + \beta_{G\times Age}(G \times Age) + \varepsilon$ | $Exp - \hat{\beta}_{G\times Age}(G \times Age)$ |
| GxSex | $Exp = \beta_0 + \beta_1 G + \beta_2 Sex + \beta_{G\times Sex}(G \times Sex) + \varepsilon$ | $Exp - \hat{\beta}_{G\times Sex}(G \times Sex)$ |
| GxTDI | $Exp = \beta_0 + \beta_1 G + \beta_2\, TDI + \beta_{G\times TDI}(G \times TDI) + \varepsilon$ | $Exp - \hat{\beta}_{G\times TDI}(G \times TDI)$ |
| GxFI | $Exp = \beta_0 + \beta_1 G + \beta_2 FI + \beta_{G\times FI}(G \times FI) + \varepsilon$ | $Exp - \hat{\beta}_{G\times FI}(G \times FI)$ |
| GxMonth[1] | $Exp = \beta_0 + \beta_1 G + \sum_{m=2}^{M} \beta_m I(Month = m) + \sum_{m=2}^{M} \beta_{G\times m}(G \times I(Month = m)) + \varepsilon$ | $Exp - \sum_{m=2}^{M} \hat{\beta}_{G\times m}(G \times I(Month = m))$ |
| GxMET[2] | $Exp = \beta_0 + \beta_1 G + \beta_2 MET + \beta_{G\times MET}(G \times MET) + \varepsilon$ | $Exp - \hat{\beta}_{G\times MET}(G \times MET)$ |
| GxE-FO[3] | $Exp = \beta_0 + \beta_1 G + \sum_{E\in e} \beta_E E + \sum_{E\in e} \beta_{G\times E}(G \times E) + \varepsilon$ | $Exp - \sum_{E\in e} \hat{\beta}_{G\times E}(G \times E)$ |
| GxE-HO[4] | $Exp = \beta_0 + \beta_1 G + \sum_{\emptyset\neq S \subseteq e} \beta_S \prod_{v\in S} v + \sum_{\emptyset\neq S \subseteq e} \beta_{G\times S}\left(G \times \prod_{v\in S} v\right) + \varepsilon$ | $Exp - \sum_{\emptyset\neq S \subseteq e} \hat{\beta}_{G\times S}\left(G \times \prod_{v\in S} v\right)$ |

[1]applied only to 25(OH)D analyses. [2]applied only to BMI analyses. [3]let e denote the set of effect modifiers included for the exposure. For LDL-C, $e = \{Age, Sex, TDI, FI\}$. For 25(OH)D, $e = \{Age, Sex, TDI, FI, Month\}$, where month is modelled as a categorical factor with M levels and represented by indicator variables I(Month=m) for m=2,…,M (with m=1 as the reference). For BMI, $e = \{Age, Sex, TDI, FI, MET\}$. [4]S denotes a non-empty subset of e, and $\prod_{v\in S} v$ denotes the corresponding interaction term among the effect modifiers in S. Exp: exposure in the falsification test (25(OH)D, BMI or LDL-C); G: genetic score for the corresponding exposure; E: effect modifier; $\hat{\beta}$: estimated regression coefficients from the model specified in the model specification column; TDI: Townsend deprivation index; FI: frailty index; GxE-FO: GxE correction with all the first-order GxE interaction terms combined; GxE-HO: GxE correction with GxE-FO plus higher-order GxE interaction terms.

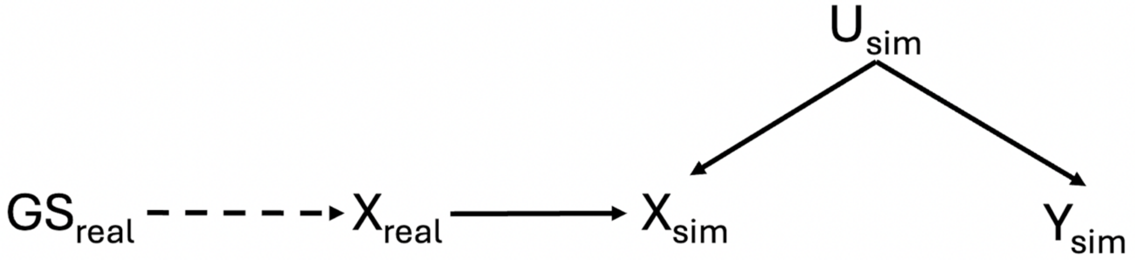


**Figure S1 Directed acyclic graph for the falsification test.** This figure is adapted from Hamilton F.W. *et al* [1]. $GS_{real}$ represents the real genetic score, and $X_{real}$ is the real exposure, downstream of $GS_{real}$. The dashed line indicates the true function that relates $GS_{real}$ to $X_{real}$ and is unknown. $U_{sim}$ is the simulated confounder. $X_{sim}$ is the simulated exposure, which is downstream of $X_{real}$ and $U_{sim}$. $Y_{sim}$ is the simulated outcome, which depends only on $U_{sim}$. There is no true causal effect of $X_{sim}$ on $Y_{sim}$. $X_{sim}$, $U_{sim}$ and $Y_{sim}$ data were simulated using the following data generating mechanism: $X_{sim} = X_{real} – U_{sim}$, $Y_{sim} = U_{sim} + e_y$, where $X_{real}$ has been scaled to have a mean of 0 and SD of 1, $U_{sim} \sim N(2, 1)$, $e_y \sim N(2, 1)$. Simulation was repeated 100 times.

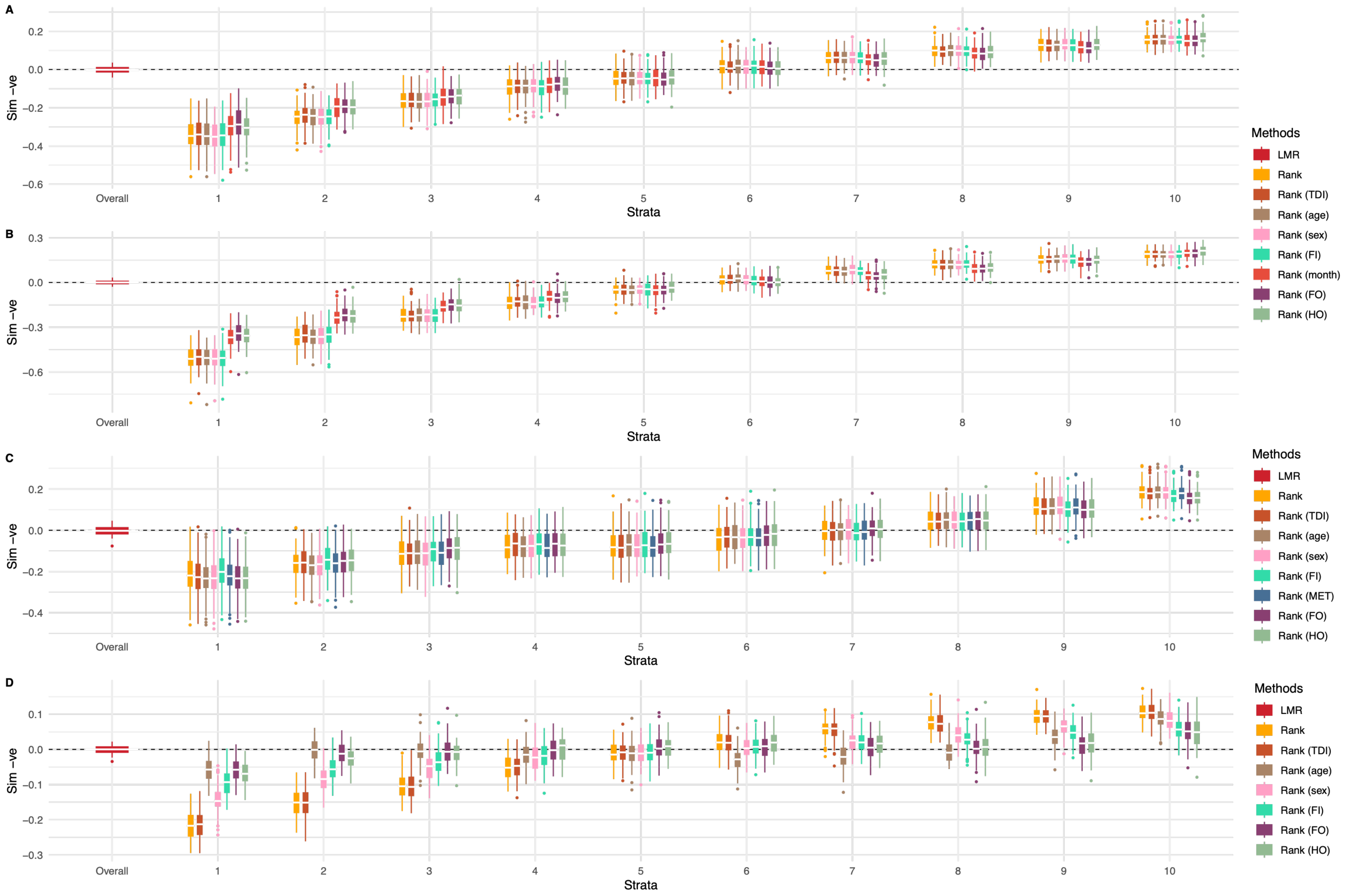
A
B
C
D
Sim −ve
Overall
1
2
3
4
5
6
7
8
9
10
Strata
Methods
LMR
Rank
Rank (TDI)
Rank (age)
Rank (sex)
Rank (FI)
Rank (month)
Rank (MET)
Rank (FO)
Rank (HO)

**Figure S2 Falsification tests for 25(OH)D using the replicated score (A), 25(OH)D using the focused score (B), BMI (C) and LDL-C (D) comparing the doubly-ranked method without GxE correction with the full set of GxE correction specifications.** The dotted line indicates the null effect. For each exposure, negative control outcome data were simulated 100 times. Boxplots represent the distribution of estimates across 100 replicates. The box displays the lower quartile, median, and upper quartile; whiskers extend to the minimum and maximum values within 1.5 x interquartile range from the lower and upper quartiles. Estimates outside this range are shown as individual points. LMR: linear MR; Rank: doubly-ranked method applied to the original exposure (no GxE correction); Rank(TDI), Rank(age), Rank(sex), Rank(FI), Rank(month) or Rank(MET): doubly-ranked method applied to a GxE-corrected exposure using a single effect modifier, namely Townsend deprivation index (TDI), age, sex, frailty index (FI), month of blood collection (month) or physical activity measured by metabolic equivalent task (MET), respectively; Rank(FO): doubly-ranked method applied to an exposure corrected using all first-order GxE interaction terms; Rank(HO): doubly-ranked method applied to an exposure corrected using the first-order GxE interaction terms plus higher-order interaction terms; TDI: Townsend deprivation index; FI: frailty index; MET: metabolic equivalent task; Sim -ve: Simulated negative control outcome.

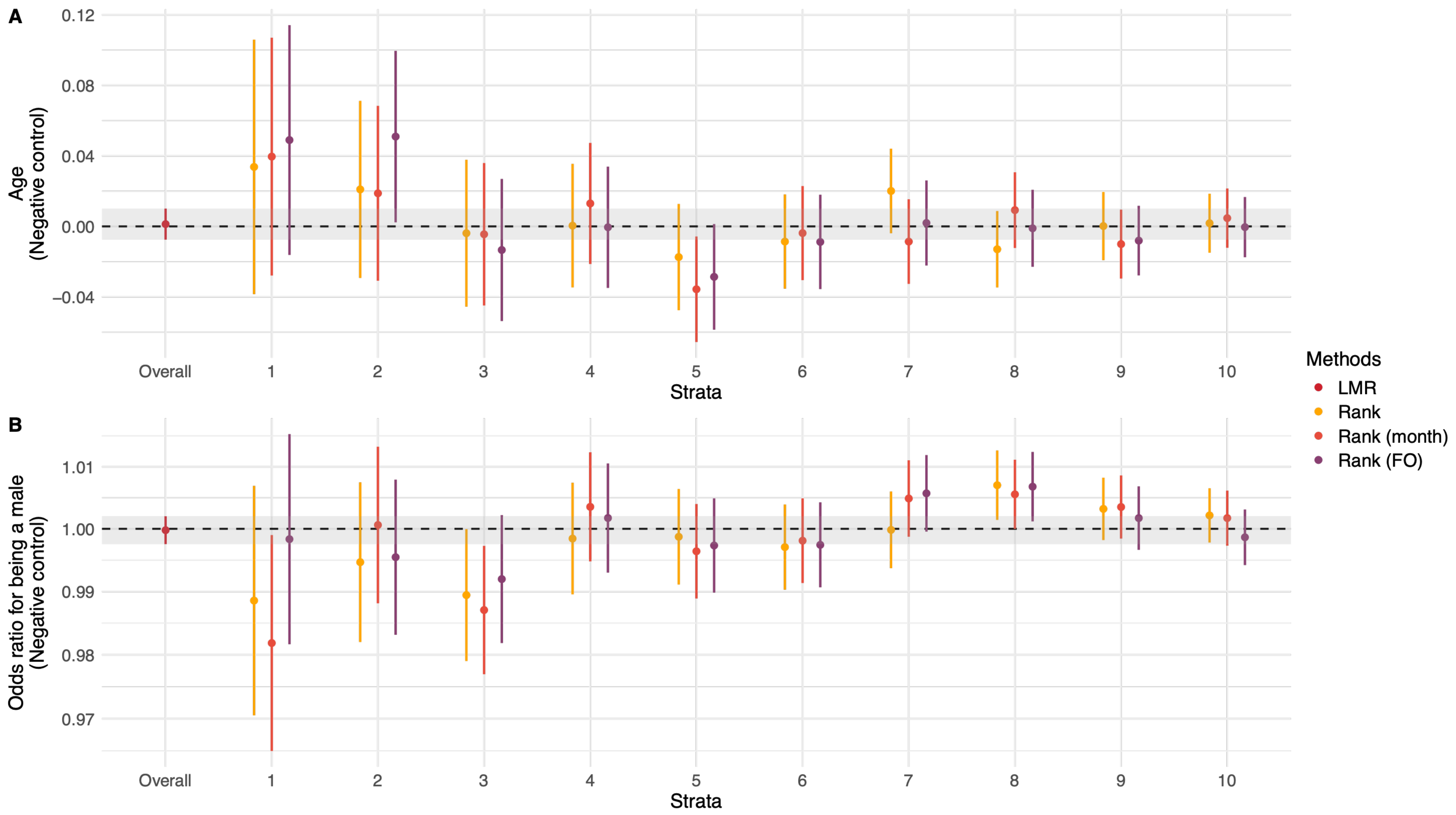


**Figure S3 Association of genetically predicted 25(OH)D using the replicated score with age at recruitment (A) and sex (B), comparing the doubly-ranked method without GxE correction with the two best-performing GxE correction specifications.** The shaded area shows the 95% CI for the linear MR estimate. The dotted line indicates the null effect. LMR: linear Mendelian randomization;

Rank: doubly-ranked method applied to the original 25(OH)D (no GxE correction); Rank(month): doubly-ranked method applied to a GxE-corrected exposure using month of blood collection (month) as the effect modifier; Rank(FO): doubly-ranked method applied to 25(OH)D corrected using all first-order GxE interaction terms. Error bars are 95% confidence intervals. When computing LACE estimates across strata, all models were adjusted for age (except when age was the outcome), sex (except when sex was the outcome), assessment centres, and top 40 genetic PCs.

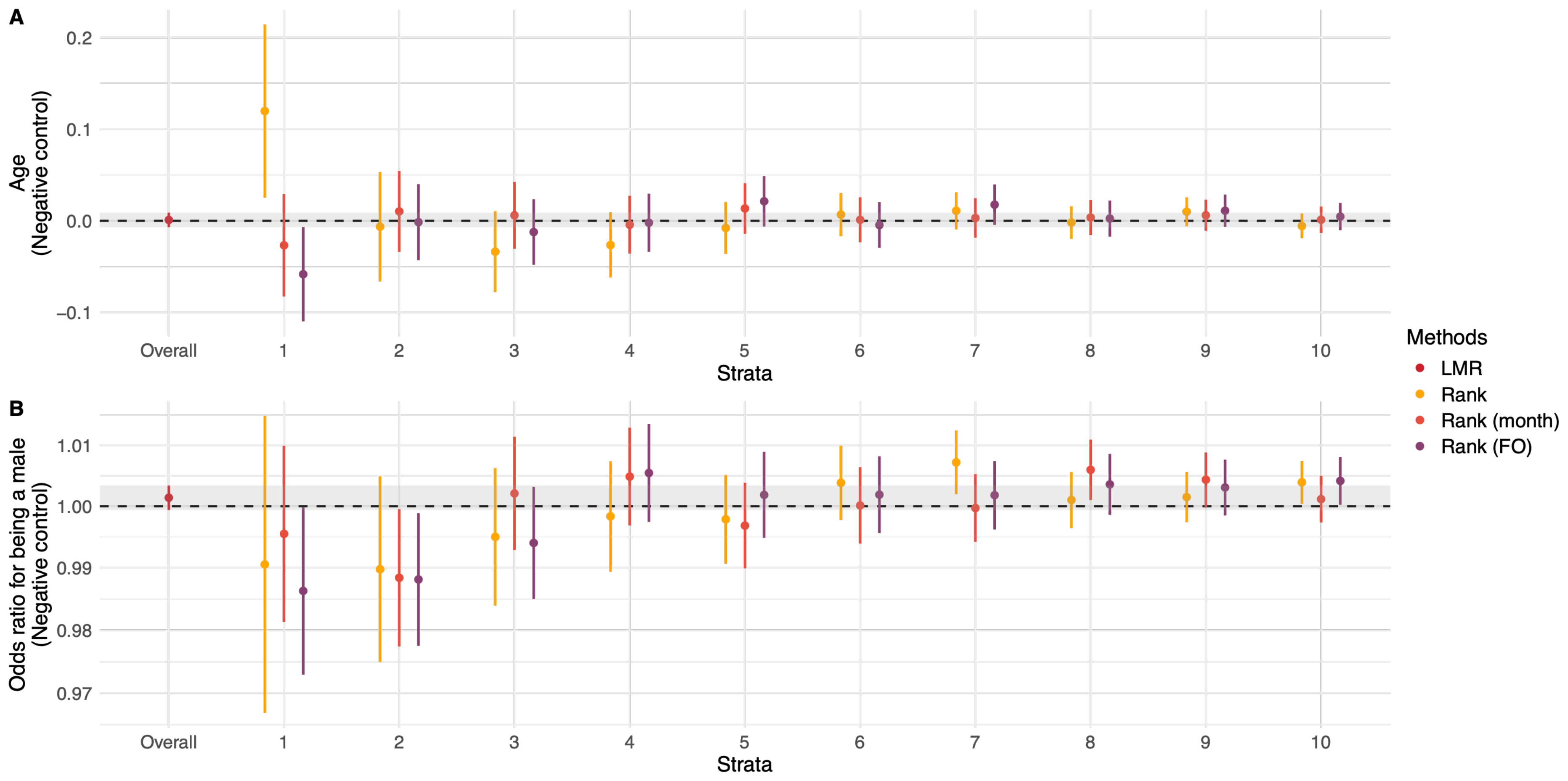


**Figure S4 Association of genetically predicted 25(OH)D using the focused score with age at recruitment (A) and sex (B), comparing the doubly-ranked method without GxE correction with the two best-performing GxE correction specifications.** The shaded area shows the 95% CI for the linear MR estimate. The dotted line indicates the null effect. LMR: linear MR; Rank: doubly-ranked method applied to the original 25(OH)D (no GxE correction); Rank(month): doubly-ranked method applied to a GxE-corrected exposure using month of blood collection (month) as the effect modifier; Rank(FO): doubly-ranked method applied to 25(OH)D corrected using all first-order GxE interaction terms. Error bars are 95% confidence intervals. When computing LACE estimates across strata, all models were adjusted for age (except when age was the outcome), sex (except when sex was the outcome), assessment centres, and top 40 genetic PCs.

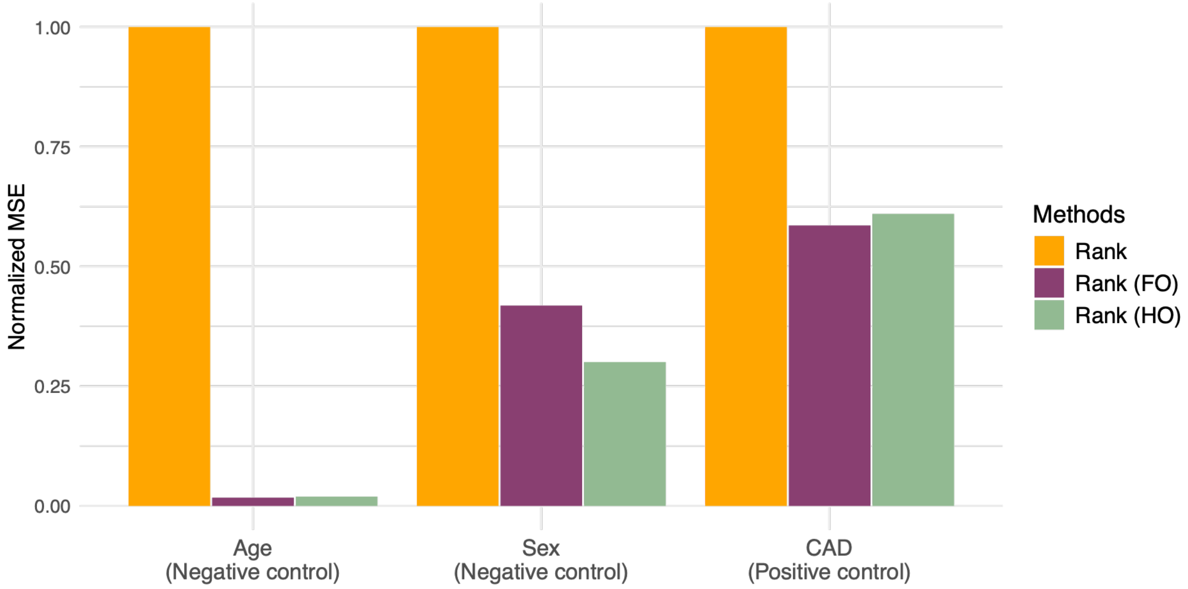


**Figure S5 Normalized mean-squared error of LACE estimates from negative and positive control analysis for LDL-C using the doubly-ranked method without and with GxE corrections.** For each method and outcome, the mean-squared error (MSE) across 10 strata was computed. For the negative controls (age at recruitment and sex), MSE was calculated as the mean of the squared LACE estimates. For the positive control (CAD), MSE was the mean of the squared differences between LACE estimates and the linear MR estimate of LDL-C on CAD risk. MSEs were normalized within each outcome by dividing by the MSE from the doubly-ranked method without GxE correction (Rank); values <1 indicate lower MSE than Rank. Rank: doubly-ranked method applied to the original LDL-C (without GxE correction); Rank(FO)[1]: doubly-ranked method applied to LDL-C corrected using all first-order GxE interaction terms; Rank(HO)[1]: doubly-ranked method applied to LDL-C corrected using the first-order GxE interaction terms plus higher-order interaction terms; CAD: Coronary artery disease. [1]Given the absence of evidence for GxTDI interaction and the negligible impact of GxTDI correction, TDI was excluded from the GxE-FO and GxE-HO correction models.
